# Prevalence and Genotypic Characterization of Extended Spectrum Beta-Lactamase Uropathogens Isolated from Refugees with Urinary Tract Infections in Nakivale Refugee Settlement camp, Southwestern Uganda

**DOI:** 10.1101/2022.04.29.22274464

**Authors:** Ayan Ahmed Hussein, Kennedy Kassaza, James Mwesigye, Bashir Mwamibi, Taseera Kabanda, Joel Bazira

## Abstract

**Background:** World Health Organization approximates that one in four individuals have had at least one UTI episode requiring treatment with an antimicrobial agent by the teen age. At Nakivale refugee camp, the overwhelming number of refugees often associated with poor living conditions such as communal bathrooms and toilets and multiple sex partners do predispose the refuges to urinary tract infections.

**Aim:** To determine the prevalence of bacterial community-onset urinary tract infections among refugees in Nakivale refugee settlement and determine the antimicrobial susceptibility patterns of the isolated pathogens.

**Methods:** This study was a cross-sectional study, that included 216 outpatients attending Nakivale Health Centre III between July and September 2020.

**Results:** Prevalence of UTI was 24.1% (52/216). The majority 86(39.81%) of the refugees were from DR Congo, followed by those from Somalia 58(26.85%). The commonest causative agent was *Staphylococcus aureus* 22/52 (42.31%) of total isolates, *followed by Escherichia coli 21/52*(40.38%). Multidrug resistant isolates accounted for 71.15% (37/52) and mono resistance was 26.92% (14/52). Out of the 52 bacterial isolates, 30 (58%) were Extended-Spectrum Beta-Lactamase organisms (ESBLs). Twenty-one (70.0%) isolates were ESBL producers while 9(30%) were non-ESBL producers. Both *bla*_TEM_ and *bla*_CTX-M_ were 62.5% each while *bla*_SHV_ detected was 37.5%.

**Conclusions:** The prevalence of UTI among refugees in Nakivale settlement is high with *Staphylococcus aureus* and *Escherichia coli as the commonest causes of UTI*. There is a high rate of multidrug resistance to common drugs used to treat UTI. The prevalence of ESBL-producing Enterobacteriaceae is high and the common ESBL genes are *bla*_TEM_ and *bla*_CTX-_

## Background

Worldwide, it has been observed that there is high dissemination of extended-spectrum beta-lactamase (ESBL)-producing uropathogens that are more severe in developing countries like Uganda [1]. Thus, ESBL producing organisms are a concern in the healthcare setting and the refugee settlement communities [2] Considerably, ESBL producing organisms often possess multidrug resistance phenotypes [3] to frequently used antibiotics to treat bacterial infections such as Beta-lactam drugs, fluoroquinolones, and aminoglycosides [4]. Besides, the widespread use of these antibiotics has caused the emergence and spread of resistant bacteria [5]. They frequently carry genes *bla*_TEM_, *bla*_CTX-_m, and *bla*sh**v** encoding resistance to aminoglycosides thus, ESBL producing organisms often possess multidrug resistance phenotypes [3]

Uganda is Africa’s largest refugee-hosting country and the 8th in the world with a total population of about 1,381,122 of which 130,462 (9.45%) are found in Nakivale refugee camp [6]. The overwhelming number of refugees is often associated with poor living conditions such as communal bathrooms and toilets and multiple sex partners resulting in compromised health [7]. As a result, a high number of suspected UTI are reported at Nakivale refugee camp (at least 20 suspected UTI per day) according to the record from the health centre.

In Uganda, studies show a high prevalence of the ESBL among patients with UTIs [8-10] Individuals dwelling in refugee settlements face numerous socio-economic, hygiene, and health care challenges [11]. These predispose to high-risk behaviors which may augment acquiring Urinary Tract Infections [12]

In Uganda, the United Nations High Commission for Refugees (UNHCR) health report indicated that 2% of the consultations registered in health centers in refugee settlements are empirically treated for Urinary Tract Infections [6]. According to unpublished health center III records in Nakivale refugee settlement, the prevalence of clinically diagnosed Urinary Tract Infections was seen to be 15%. Resistant Urinary Tract Infections are among the challenges faced by health care providers in refugee population[13]. Accordingly, therapy is empiric and with the influx of refugees, new infections and uropathogenic gram negative bacterial resistant strains could have been introduced as already reported by some studies [14]. In such cases, such resistant strains can easily spread in the refuge settlement camps and delayed recognition of severe UTIs caused by ESBL producers, and inappropriate treatment with antibiotics has been associated with increased mortality and morbidity in resettlement camps a phenomenon that stresses the need for evaluation of Urinary Tract Infections etiology.

## Materials and Methods

### Study design, duration, and site

The study was a descriptive cross-sectional study, conducted among 216 outpatients attending Nakivale Health Centre III in Nakivale Health Centre III refugee settlement, Isingiro district, Southwestern from July 2020 to September 2020.

### Inclusion and Exclusion criteria

The study enrolled participants with complaints suggestive of UTI such as dysuria, frequency, urgency, suprapubic pain, cloudy or malodorous urine, or flank pain, fever as identified by the attending clinician and who were 18 years, and above were considered after signing informed consent. We excluded participants who failed to provide a urine sample, female who were in their menstruation period.

### Data collection procedure

Participants were recruited from Nakivale Health Centre III. This health facility was selected considering that it is the main health center offering services to the refugee community in Nakivale refugee settlement. Eligible consented participants were administered with a questionnaire translated into the local language i.e Kiswahili, Somali, Kinyarwanda. Information regarding the participants’ socio-demographics was obtained directly by interviewing the participants. This process was facilitated by research assistants who understand the language, and they were also trained on the questionnaire before the commencement of the study. Further, the participant’s clinical presentation was obtained from the Outpatient department. Data collected included but was not limited to; age, sex, marital status, education status, country of origin, comorbid illnesses (diabetes), history of UTI, antibiotics administered in the past month. Besides, a urine sample was collected and sent to the laboratory for visual reporting, microscopic wet preparation, biochemical, culture, and antibiotic susceptibility testing.

### Specimen collection and transportation

Study participants were given sterile urine containers that are leakproof. The containers were labelled very well with the study participant’s details (Study Number, date, and time of sample collection). The participants were instructed on how to collect the MSU sample, (to avoid contamination of the specimen, all participants were told to first cleanse the urethral and labia area with clean water) and then collected the MSU into a wide mouth clean sterile container. These samples were then packed into a cool box (2-8 degree centigrade) and transported to Mbarara University Microbiology laboratory within 4 hours.

proper instructions in form of written and oral were given to the participants on the collection of midstream urine with minimum contamination.

### Laboratory tests Urine culture and isolation of Organisms

The urine samples were received and analyzed at the microbiology laboratory of MUST. The appearance (color and turbidity) of the urine samples were recorded. Microscopic wet-preparation of the uncentrifuged urine was done to examine significant pyuria (>10 cells/µl), casts, red cells, bacteria, and yeast. Also, biochemical testing using a urine dipstick was done to detect protein, nitrite, and leukocyte esterase. Then the urine was analyzed using a dipstick, and findings were reported in the register. Also, primary Gram staining was done. Further, the urine samples were inoculated using a standard loop by transferring 5µL of urine and plated on Cystine Lactose Electrolyte-Deficient (CLED) agar and incubated aerobically at 37°C overnight for 48 hours. The following day, for such samples that had grown on the media, colonies were counted 50 and above colony forming units which were equivalent to 10000 (10^4^) by ml (CFU/mL) and was used to determine the significance of the growth of Positive UTI[15]. This was used to calculate the prevalence of UTI 24.1%.

The bacteria etiology causing UTI were identified from those that showed significant growth. These were purified on Macconkey Agar, Blood Agar, and mannitol salt Agar. for further identification using biochemical tests and Analytical profile index (API).

### Biochemical tests

Biochemical tests were carried out after performing Gram staining. For Gram-Negative bacilli/ Cocco-bacilli, the organisms were differentiated into Lactose fermenters and non-lactose fermenters. Lactose fermenters were subjected to different panels of biochemical tests. These tests included TSI, Indole, Urea, Motility, Methyl Red/ Vogues prosker, and citrate fermentation while non-lactose fermenters were tested for Glucose oxidation and enzyme oxidase production and later differentiated into, oxidase-positive and Oxidase Negative.

Gram-positive cocci were tested for catalase enzyme production with 3% Hydrogen peroxide. Catalase positive were later subjected to anaerobic conditions, glucose fermentation, and coagulase and DNAse enzyme tests. On the other hand, Catalase negative organisms were subjected to Bacitracin sensitivity, Hippurate hydrolysis, bile-esculin hydrolysis, and Tolerance to 6.5% NaCl.

### Analytical profile index (API) tests

For definitive identification of *Escherichia coli, Klebsiella pneumoniae, and Salmonella arizonae, Citrobacter freundii, Hafnia spp*, we used API20E

### Antimicrobial susceptibility testing

The antimicrobial susceptibility pattern of the isolated bacterial pathogens was performed using the disc diffusion method on Mueller-Hinton agar according to the guidelines of clinical and laboratory standards institute. For this, a few discrete from an 18 to 24 hour-old culture colonies were picked using a sterile inoculation loop and emulsified in sterile peptone water to form a suspension by vortexing. The turbidity of the suspension was adjusted visually to match that of the 0.5 McFarland standard. the suspension was evenly distributed over the entire surface of well-dried Muller Hinton agar plates using a sterile cotton-tipped swab. Using sterile forceps, the antibiotic discs were placed on the inoculated plates and incubated at 37°C for 24 hours. The antibiotic discs and their relative potency that were used include; Nitrofurantoin (100 µg), Ampicillin (10µg), Ciprofloxacin (5µg), Ceftriaxone (30µg), Amoxicillin-clavulanic acid (20/10µg), Erythromycin (15µg), Chloramphenicol (30µg), Penicillin (10µg). The diameter of the zone of inhibition around the disc was measured using a digital metal caliper and isolates were classified as sensitive, intermediate, and resistant according to Clinical and Laboratory Standards Institute guidelines [16]. To maximize safety, handling and processing of urine samples were done using personal protective equipment like gloves, goggles, face masks, and laboratory coats. Universal safety precaution was followed at all times. Resistance was then categorized into Monoresistance (Mono) if there was insusceptibility to one of two classes of antimicrobials or Multidrug resistance (MDR) if there was insusceptibility to three (3) or more antibiotics and extremely resistant (XDR) if there was insusceptibility to all antibiotics tested

### Screening for ESBLs

The screening test is based on testing the organism for resistance to an indicator cephalosporin. is commonly used as it is hydrolyzed by TEM, SHV, and CTX-M types, but other cephalosporins such as cefotaxime, cefotaxime-clavulanic acid (CTC), and ceftazidime, ceftazidime-clavulanic acid (CZC) and are also used following Clinical and Laboratory Standards Institute recommendations. The use of more than one of these agents for screening improves the sensitivity of detection [17]

### Phenotypic disc confirmatory test (PDCT)

There are several commercial tools used to confirm these Enterobacteriaceae suspensions were adjusted to 0.5 McFarland standard organisms, including double-disc synergy, combination disc method [18]. Phenotypic confirmatory disc diffusion tests are done on the zone diameters that indicate suspicion for ESBL production, thus it helps to confirm if the isolates are ESBL producers or not. The drugs used in double-disc synergy include; *Ceftazidime (CAZ)* disc containing 30μg of the antibiotic and ceftazidime-clavulanic acid (CZC) disc containing 30/10μg of the antibiotics and *cefotaxime (CTX)* disc containing 30μg of the antibiotic and a cefotaxime-clavulanic acid (CTC) disc containing 30/10μg of the antibiotics [19].

These antibiotics are placed at a distance of 30 mm apart, the plates are incubated overnight at 37°C in ambient air and the results for inhibition zones are read. A ≥ 5 mm increase in the zone diameter for CZC, versus its zone diameter, when it tested alone by CAZ and/or a ≥ 5 mm increase in the zone diameter for CTC, versus its zone diameter when it tested alone by CTX, was used to confirm presence ESBL as recommended by clinical laboratory standards institute guidelines. *Klebsiella pneumoniae* ATCC 700603 was used as an ESBLs-positive reference strain while Escherichia *coli* ATCC 25922 was used as an ESBL-negative reference strain [16, 20]

### Genotypic characterization of ESBLs

#### Genomic DNA extraction

Isolates were sub-cultured at 37°C overnight in Muller Hinton Agar (Oxoid, Wade Road, Basingstoke, UK). The bacterial DNA was extracted using the Zymo-Research quick-*g*-DNA Miniprep kit and protocol, v1.0.0. Briefly, 400uL of Zymo kit Lysis buffer was dispensed into a 1.5mL conical tube using a P1000 micropipette. A loopful of bacterial colonies from the overnight culture was emulsified in the buffer. The tubes were vortexed for approximately 10 -15 seconds and allowed to stand for 10 minutes. The entire content of the tube was transferred into the Zymo Research Spin column, placed inside a collection tube, and centrifuged at 10,000rpm for 1 minute.

The collection tube together with the filtrate was discarded and the spin column was placed in a new collection tube and 200uL of gDNA pre-wash buffer was added into the spin column and the assembly was again centrifuged for 1 minute at 10,000rpm. The collection tube and content were discarded and the spin column was placed into a new collection tube. To each Spin column in a collection tube, 500uL of gDNA Wash buffer was added and the assembly centrifuged for 3 minutes at 10,000rpm. The collection tube and its content were discarded and the mini-column placed into a clean, newly labeled 1.5mL tube. For final DNA elution, 50uL of the elution buffer was added into the spin column and allowed to incubate at room temperature for 5 minutes. The column-tube assembly was centrifuged at 10000rpm for 1 minute and the DNA collected was stored at -20**°C** before use.

### Characterization of ESBL genes using PCR

The presence of genes encoding ESBL was detected using PCR amplification. This study detected bla-SHV, bla-CTX-M [21], and bla-TEM [22], genes using primers from the previous studies. The PCR master mix reagents for each gene target were as follows: 12.5 µL master mix consisting of One Taq quick-load 2x master mix/w standard buffer, dNTPs & Taq polymerase (M0486S), 1.5 µL forward (100 µM), 1.5 µL primary reverse (100 µM), 5 µL DNA template and RNAase-free water up to 25 µL.

#### PCR Cycling

The PCR process was carried out in a thermocycler (Multigene Optimax) with a pre-denaturation cycle of 95°C for 15 min, followed by DNA amplification stage with 30 cycles (94°C for 1 min, 54.5-60°C for 1 min, and 72°C for 1 min) and final extension cycle of 72°C for 5 min.

#### Gel electrophoresis

DNA Amplicon was electrophoresed using 1.5% agarose gel, in Tris-Borate EDTA buffer (TBE) 1×concentration, Safe View Classic™ DNA stain (cat # G108), 6x loading dye (Thermo Scientific #R0611), and DNA ladder/marker 100 bp (NEB-Biolabs #N3231L). DNA Bands were visualized on a Dark reader Transilluminator (DR46-B).

### Quality control

The study incorporated quality control checks to ensure reliable processing. In this, the emphasis was on strict adherence to standard operating procedures on sample collection, transport, and sample analysis. There was the matching of sample labels against the accompanying laboratory forms to ensure that no mix-up occurs. Culture media was made according to manufactures instruction, and this was tested for sterility, and performance tests (fertility check) were done. Reference strains of *E. coli* ATCC25922, *Klebsiella pneumoniae* ATCC 700603, and *S. aureus* ATCC 29213. *Pseudomonas aeruginosa* ATCC 27853, *Enterococcus faecalis* ATCC 29212 were used for the quality control of the antibiotic discs and extended-spectrum beta-lactamase (ESBLs) [20]

### Data management and analysis

The obtained data were entered and cleaned in Microsoft Excel and, and was imported to STATA version 12.0 for analysis. Evaluations were carried out at 95 % confidence level (95% CI) and values of (p<0.05) were regarded as significant. Results were presented in form of tables, pie charts and graphs.

### Ethical consideration

This is study was approved by Research and ethics committee of Mbarara University of Science and Technology. Upon clearance, permission was sought from the Office of the Prime Minister to research a refugee population, and informed consent was sought from each participant. All methods were performed in accordance with the relevant guidelines and regulations. The participants who had UTIs were referred to clinicians for further management.

### Data Availability

The datasets used and/or analyzed during the current study available from the corresponding author on reasonable request.

## RESULTS

### The demographic characteristics of study participants

The study recruited 216 clients among whom 164 (75.93%) were females and 52 (24.07%) were males. The age range of participants was 18 to 76 years with an average age of 35.86 (SD=11.218) and a median age of 34 years. The majority 86(39.81%) of the refugees were from DR Congo, followed by those from Somalia 58(26.85%). Regarding education many 89(41.20%) were not educated, followed by those primary level accounting to 82(37.96%). The detailed findings are shown in table 2 below.

**Table 1:**
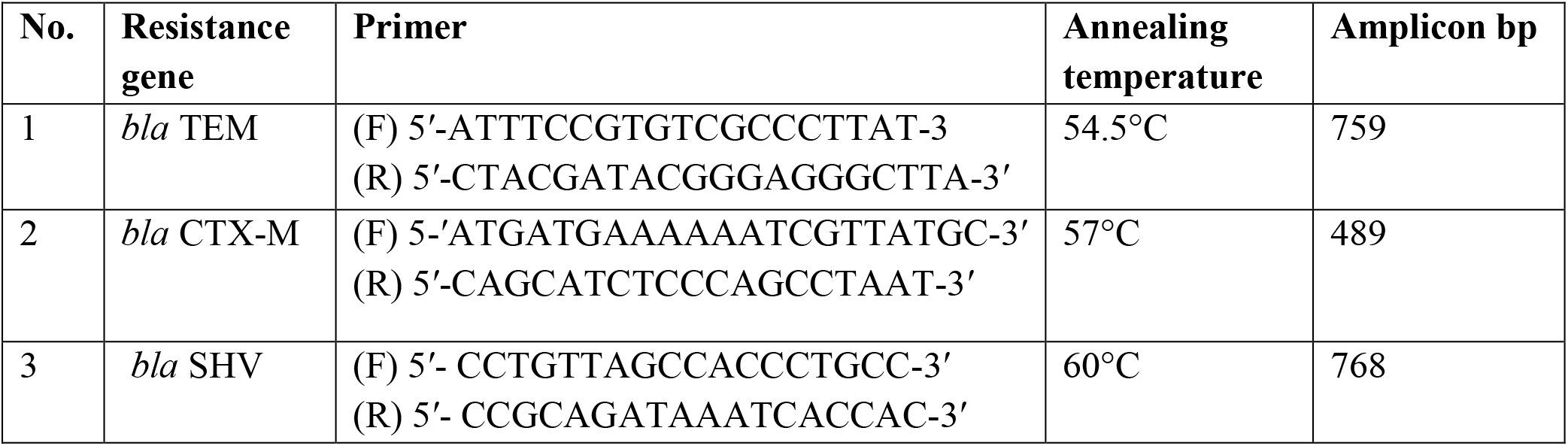
Primers sets used in the detection of ESBL encoding genes from the study isolates[23].

**Table 2:** Demographic characteristics of the study participants.

| N=216 |  |  |
| --- | --- | --- |
| Variable | Frequency (n) | Percentage (%) |
| **Sex** |  |  |
| Female | 164 | 75.93 |
| Males | 52 | 24.07 |
| **Age group** |  |  |
| 18-76 years | 216 | 100 |
| **Country of origin** |  |  |
| Burundi | 27 | 12.50 |
| DRC | 84 | 38.89 |
| Ethiopia | 01 | 0.46 |
| Rwanda | 35 | 16.20 |
| South Sudan | 11 | 5.09 |
| Somalia | 58 | 26.85 |
| **Education level** |  |  |
| None | 89 | 41.20 |
| Primary | 82 | 37.96 |
| Secondary | 37 | 17.13 |
| Tertiary | 8 | 3.70 |
| **Marital status** |  |  |
| Divorced | 15 | 6.94 |
| Married | 163 | 75.46 |
| Single | 27 | 12.50 |
| Widowed | 11 | 5.10 |

### The prevalence of urinary tract infections among refugees in Nakivale refugee settlement

Out of the 216 samples collected and cultured, 52 had significant growth of bacteria while 164 showed no growth. The overall prevalence of UTI in this study was 24.1%.

### Prevalence of UTI based demographic factors

The prevalence of UTI was 25.00% (41/164) among females and 21.15% (11/52) among males. With country-of-origin South Sudan had the highest prevalence of UTI which was 36.36% (4/11), followed by DRC 29.76% (25/84). Refugees of primary level of education had the highest prevalence of UTI 29.27% (24/84), followed by uneducated 23.60% (21/89). Lastly, single refugees had the highest prevalence of UTI 29.63% (8/27), followed by married refugees 23.93% (39/163). (see details in table 3)

**Table 3:** Distribution of UTI prevalence by sex.

| **Sex** | **Positive cultures for bacteria (52)** | **Prevalence of UTI** |
| --- | --- | --- |
| Female (164) | 41 | 25.00% |
| Male (52) | 11 | 21.15% |
| **Country of origin** |  |  |
| Burundi (27) | 7 | 25.93% |
| DRC (84) | 25 | 29.76% |
| Ethiopia (1) | 0 | 0.00% |
| Rwanda (35) | 5 | 22.73% |
| South Sudan (11) | 4 | 36.36% |
| Somalia (58) | 11 | 18.96% |
| **Education level** |  |  |
| None (89) | 21 | 23.60% |
| Primary (82) | 24 | 29.27% |
| Secondary (37) | 6 | 16.22% |
| Tertiary (8) | 1 | 12.50% |
| **Marital status** |  |  |
| Divorced (15) | 3 | 20.00% |
| Married (163) | 39 | 23.93% |
| Single (27) | 8 | 29.63% |
| Widowed (11) | 2 | 18.18% |

### The bacterial etiology of urinary tract infections among refugees in Nakivale settlement

The majority of the organism was *Staphylococcus aureus* accounting for 22/52 (42.31%) of total isolates, *followed by Escherichia coli 21/52*(40.38%), *Klebsiella pneumoniae 4/52*(7.69%), *Salmonella arizonae 2/52*(3.85%) *while Citrobacter freundii, Hafnia spp, and Pseudomonas aeruginosa* accounted to 1/52(1.92%) each as illustrated in in figure 1

**Figure 1:**
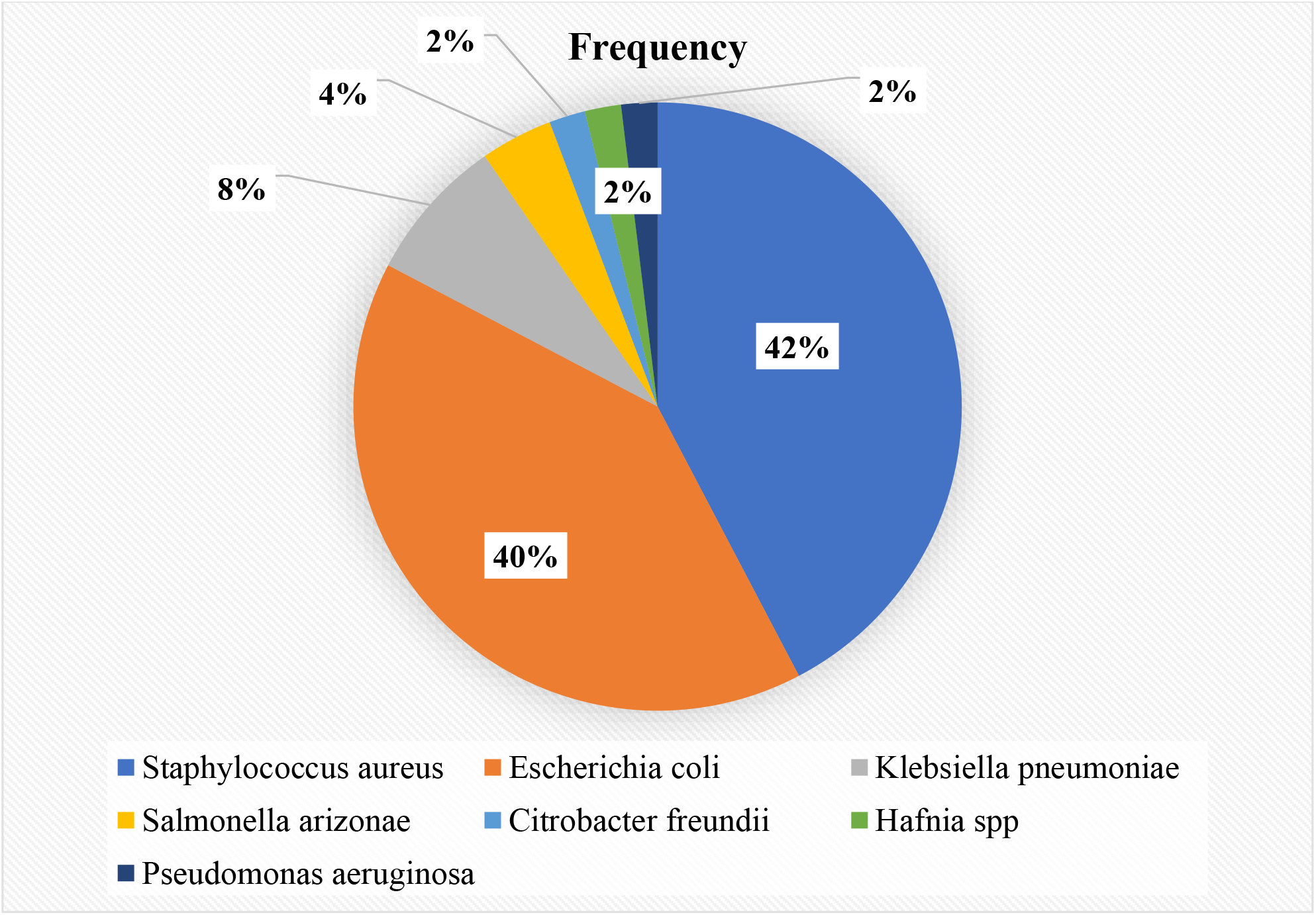
The bacterial etiology of urinary tract infections among refugees in Nakivale settlement.

### The antimicrobial susceptibility pattern of the isolated bacterial pathogens among refugees in Nakivale settlement

According to table 4 below, the majority of the isolates showed resistance to at least one class of antimicrobials that is many 37(71.15%) of the bacteria isolates were multi-resistant, 14(26.92%) were mono-resistant to anti-biotics, and last 1 (1.92%) bacteria isolate was not resistant to any antibiotic.

**Table 4:** The antimicrobial susceptibility pattern of the isolated bacterial pathogens among refugees in Nakivale settlement

| Resistance classification | Frequency | Percentage |
| --- | --- | --- |
| None | 1 | 1.92 |
| Mono | 14 | 26.92 |
| MDR | 37 | 71.15 |
| Total | 52 | 100.00 |

#### 4.4.1 Antimicrobial susceptibility pattern of the individual isolated bacterial pathogens among refugees in Nakivale settlement

*Klebsiella pneumoniae, Salmonella arizonae, Citrobacter freundii, Hafnia spp, and Pseudomonas aeruginosa*, isolates were multidrug-resistant. All *E. coli* isolates were resistant to at least one class of antimicrobials that are 33.33% being mono-drug-resistant and 66.67% being multidrug-resistant. Lastly of the 22 *S*.*aureus* isolates only 1 (4.55%), the isolate was not resistant to any antimicrobial as in the table 5 below;

**Table 5:** Antimicrobial susceptibility pattern of the individual isolated bacterial pathogens among refugees in Nakivale settlement

| Isolates | Non-drug-Resistance<br>No. (%) | Mono-Drug-Resistance<br>No. (%) | Multidrug-Resistance<br>No. (%) | Total (%) |
| --- | --- | --- | --- | --- |
| *S.aureus* | 1(4.55) | 7(31.82) | 14(63.63%) | 22 (100%) |
| *E. Coli* | 00.00 | 7(33.33) | 14(66.67) | 21 (100%) |
| *Klebsiella pneumoniae* | 00.00 | 00.00 | 4(100%) | 4 (100%) |
| *Salmonella arizonae* | 00.00 | 00.00 | 2(100%) | 2 (100%) |
| *Citrobacter freundii* | 00.00 | 00.00 | 1(100%) | 1 (100%) |
| *Hafnia spp* | 00.00 | 00.00 | 1(100%) | 1 (100%) |
| *Pseudomonas aeruginosa* | 00.00 | 00.00 | 1(100%) | 1 (100%) |

### Prevalence of Extended Spectrum Beta-Lactamase organisms ESBLs among the organisms isolated in refugees at Nakivale refugee settlement

Out of the 52 bacteria isolates, 30 (58%) were Extended-Spectrum Beta-Lactamase organisms (ESBLs) while 22 (42%) were none ESBLs. Therefore, the prevalence of Extended Spectrum Beta-Lactamase organisms ESBLs in this study was 58%.

### Phenotypic characterization of Extended Spectrum Beta-Lactamase organisms ESBLs among refugees in Nakivale refugee settlement

Phenotypic characterization of ESBL production was performed on 30 Gram-negative organisms and 21 (70.00%) were ESBL producers for the different organisms while 9(30%) were Non-ESBL producers as indicated in table 6 below.

**Table 6:** ESBL production by the different organisms.

| **ORGANISMS**<br>**ISOLATED** | **ESBL PRODUCER** |  | **TOTAL** |
| --- | --- | --- | --- |
|  | **NO (%)** | **YES (%)** |  |
| Escherichia coli | 7(33.33) | 14(66.67) | 21 |
| Klebsiella pneumoniae | 1(25.00) | 3(75.00) | 4 |
| Salmonella arizonae | 1(50.00) | 1(50.00) | 2 |
| Citrobacter freundii | 0(0.00) | 1(100%) | 1 |
| Hafnia spp | 0(0.00) | 1(100) | 1 |
| Pseudomonas aeruginosa | 0(0.00) | 1(100) | 1 |
| **TOTAL** | **9(30.00)** | **21(70.00)** | **30** |

**Genotypic characterization of Extended Spectrum Beta-Lactamase organisms ESBLs among refugees in Nakivale refuge**out of 30 Extended-Spectrum Beta-Lactamase organisms, the gene was detected in 8(26.67%). Of which the most common genes were *bla*_TEM_ and *bla*_CTX-M_ each detected in 5 (62.5%) of the organisms with genes. While ***bla***_**SHV**_ was detected in 3 (37.5%) of the organisms with genes as shown in table 7 below.

**Table 7:**
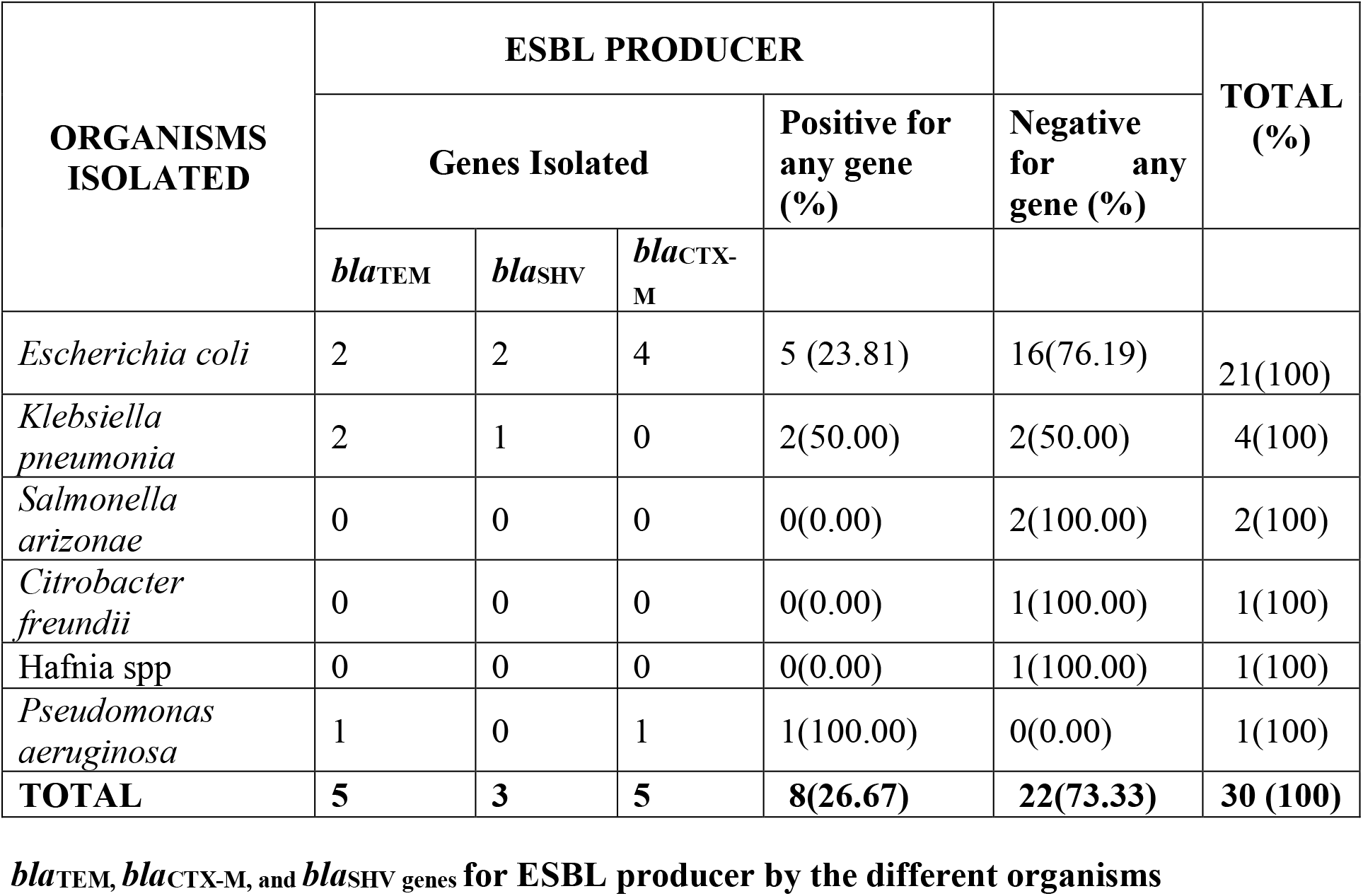
Genotypic characterization of Extended Spectrum Beta-Lactamase organisms.

| ORGANISMS<br>ISOLATED | ESBL PRODUCER |  |  |  |  | TOTAL<br>(%) |
| --- | --- | --- | --- | --- | --- | --- |
|  | Genes Isolated |  |  | Positive for<br>any gene<br>(%) | Negative<br>for any<br>gene (%) |  |
|  | *bla*<sub>TEM</sub> | *bla*<sub>SHV</sub> | *bla*<sub>CTX-M</sub> |  |  |  |
| *Escherichia coli* | 2 | 2 | 4 | 5 (23.81) | 16(76.19) | 21(100) |
| *Klebsiella pneumonia* | 2 | 1 | 0 | 2(50.00) | 2(50.00) | 4(100) |
| *Salmonella arizonae* | 0 | 0 | 0 | 0(0.00) | 2(100.00) | 2(100) |
| *Citrobacter freundii* | 0 | 0 | 0 | 0(0.00) | 1(100.00) | 1(100) |
| *Hafnia* spp | 0 | 0 | 0 | 0(0.00) | 1(100.00) | 1(100) |
| *Pseudomonas aeruginosa* | 1 | 0 | 1 | 1(100.00) | 0(0.00) | 1(100) |
| **TOTAL** | **5** | **3** | **5** | **8(26.67)** | **22(73.33)** | **30 (100)** |

*bla*<sub>TEM</sub>, *bla*<sub>CTX-M</sub>, and *bla*<sub>SHV</sub> genes for ESBL producer by the different organisms

## DISCUSSION

### The prevalence of urinary tract infections

In this study, the prevalence of UTI was 24.1%. This finding was consistent with the finding in a study done in Bushenyi by Tibyangye [24] where the prevalence of UTI was (22.5%) and the prevalence in a study done in Gulu by Odongo [25] which was (24.2%). Therefore, the reason for the consistent results could behave resulted from geographic locations as suggested by Tandogdu, since all study was in Uganda regardless of having the study population as refugees.

The finding was high when compared 16.3% in a study done among refugees in Turkey by Turktan 13.3% in a study done among refugees in Finland by Aro [28], 5% in a study done among Syrian refugees in Asia by Gulati [29] and 7% in Tanzania as showed by UNHCR [30]. Aaccording to Uganda refugee health report 2018, out of all consultations made in all refugee serving health facilities in the refugee settlements, UTI comprised 2%, a figure contradicting this study found. Thus, the reason for this differing prevalence of UTI could due to the methods used to diagnose UTI. In our study, we used urine culture whereas they used simple urinalysis.

However, the study prevalence was low when compared to 38.8% in a study done at Mulago hospital in Uganda, by Kabugo [31] and (32.2%) in a study done among patients attending hospitals in Bushenyi [32]. Although our prevalence is not significantly lower than the ones from Mulago hospital and KIU teaching hospital, is bevies our study recruited mostly outpatients who came to lower health units as compared to patients that seek care at a tertiary hospital.

The reason for the high prevalence of UTI could be due to the overwhelming number of refugees which is often associated with poor living conditions such as communal bathrooms and toilets and multiple sex partners resulting in compromised health[7]

### The bacterial etiology of urinary tract infections

Although other studies have found that *E. coli* was the most common organism causing UTI, [33, 34] in our study the commonest organism was *S. aureus*. In this study, the most common bacterial isolates were *Staphylococcus aureus* accounting for 42.31% of total isolates, *followed by Escherichia coli* (40.38%). This was consistent with a study done in Nigeria by Ekwealor [35] who found that *S. aureus* (28%), was the common isolate, followed by *E. coli* (24.6%). Also results in a study was done by Regea [36] in Ethiopia found that *S. aureus* were the most common isolates accounting for (24.2%), *CoN Staphylococcus spp* (24.2%), *E. coli* (12.1%), and *K. pneumonia* (12.1%).

### The antimicrobial susceptibility pattern of the isolated bacterial pathogens

In this study, many 37(71.15%) of the bacteria isolates were multi-resistant, 14(26.92%) were mono-resistant to anti-biotics, and one (1.92%) bacterial isolate was not resistant to any antibiotic. This finding is low when compared to 89.5% MDR in the study done in Ghana by Agyepong [37], 78% of the isolates were multi-drug resistant (MDR) in 2014 in a study done in Netherlands[38] and 83% of the drugs that were multidrug-resistant to bacteria isolates according to Manikandan [39]

However, it was high when compared to 23% of the isolates were multi-drug resistant (MDR) in 2017 in a study done in Netherlands by Stalenhoef [38], 63% of the isolates that were multidrug-resistant (MDR) in a study done by Ramirez-Castillo [40], 54.2% of the bacterial isolates were multi-drug resistant in a study done in Bangladesh [41], 41.1% isolates that were multi-drug resistant in a study done in Nepal by Baral [42], 61.4% of the *bacteria* isolates that were multidrug-resistant in a study done in Egypt [43] and lastly 14.5% multidrug-resistance rate in the study done in the south and eastern Europe, Turkey and Israel by [44]. This high finding of multidrug resistance rate in this study could be because the most dominant bacteria isolate (*E. coli* in this study) has shown a substantial multidrug resistance to antibiotics normally given for the management of UTIs in studies by Odongo[45], and [46]

### Antimicrobial susceptibility pattern of the individual isolated bacterial pathogens among refugees in Nakivale settlement

In this study all *Klebsiella pneumoniae, and Pseudomonas aeruginosa*, isolates were multidrug-resistant. This finding was high when compared to results in a study done in the south and eastern Europe, Turkey, and Israel which found that the highest multidrug-resistance was (54.2%) *Klebsiella pneumoniae* and followed by 38.1% multidrug resistance rate for *Pseudomonas aeruginosa* according to Gomila [44].

In this study 66.67% *E. coli* and all the *Citrobacter freundii*, isolates were multidrug-resistant. This finding low when compared to results in a study done in Nepal [47] which found that *E. coli* isolates were 38.2% MDR, and *Citrobacter spp*. was 72.7% MDR.

However, the results similar to those in a study done in Ghana [37] since both studies showed that all (100%) *P. aeruginosa* were multidrug-resistant. Therefore, there is a high multidrug resistance rate of individual bacteria isolates to antibiotics used to treat UTIs.

### Prevalence of Extended Spectrum Beta-Lactamase organisms ESBLs among the organisms isolated

In this study, the prevalence of Extended Spectrum Beta-Lactamase organisms ESBLs among organisms isolated was 58% of the 52 isolates. This was low when compared to the prevalence of 90.16% in a study conducted by Shrestha [48] in Sahid Memorial Hospital, Kalanki, Nepal from August 2014 to January 2015, the prevalence of 86.67% of 225 isolates in a study done in India by Aruna and Mobashshera, [49] and the prevalence 68.60% of 86 isolates in a study conducted by Odoki [33] in Bushenyi, Uganda. Therefore, the reason for the low prevalence of ESBLs in this could have resulted from low bacteria isolated found.

However, it was lower than the prevalence of 47.69% of 216 bacteria isolate in a study done by Ekwealor [35] in Nigeria which weakens low bacteria isolated as a reason for the low prevalence of ESBLs in this study. Thus, the prevalence of Extended Spectrum Beta-Lactamase organisms ESBLs among organisms isolated was low in this study.

### Phenotypic characterization of Extended Spectrum Beta-Lactamase organisms ESBLs

In this study phenotypic characterization of ESBL production was performed on 30 Gram-negative organisms and 21 (70.00%) were ESBL producers for the different organisms while 9(30%) were non-ESBL producers. This prevalence of ESBL was high when compared with the overall prevalence of ESBL was 25%, in Africa 33%, in India 15%, North and south America was 3%, in other Asian countries it was 5% and in Europe, it was 4% according to a study carried out by (Mansouri et al., 2019). Also, higher than 55.5% prevalence of ESBL positive organisms in the study done by Magale [50], in Nairobi Kenya, 56% in a study done by Kasango [51] in Uganda, and 64.9% in a study done by Kateregga [52] in Uganda.

Therefore, this high prevalence of ESBL-producing Enterobacteriaceae reflects a more serious public health problem since according to Adler [53] dissemination of ESBLs compromises the activity of broad-spectrum antibiotics creating major therapeutic difficulties with a significant impact on the outcomes for patients. To clarify this a study by (Fatima et al., 2018) showed that 97% of ESBL producing bacteria isolates were resistant to Ampicillin. Thus, Ampicillin resistance in this study could have resulted from the high prevalence of ESBL producing Enterobacteriaceae in this study.

Also, according to results in a study by Sakina [54], ESBL producing bacteria isolates were 59% resistant to Ciprofloxacin and 50% resistant to Ceftriaxone. Therefore, the high prevalence of ESBL producing bacteria isolates could also be the reason for the high multi drug resistance in this study.

### Genotypic characterization of Extended Spectrum Beta-Lactamase organisms ESBLs

In this study, the most common genes were *bla*_TEM_ and *bla*_CTX-M_ each detected in 5 (62.5%) of the organisms with genes. While ***bla***_**SHV**_ was detected in 3 (37.5%) of the organisms with genes. This contradicts the finding in studies done by Jena [55, 56] wherein all these studies *bla*_TEM_ the only predominant gene was followed by *bla*_CTX-M_. Other contradictions were reflected by studies conducted by Seyedjavadi [57] and Ahmed [58] were for their case *bla*_CTX-M_ was the predominant gene was followed by *bla*_TEM._ The reason for such could be the difference in study areas and people.

Despite the difference on which gene is common the percentage of *bla*_TEM_ and *bla*_CTX-M_ in this study and others was above 60%. Thus, it can be conclusive that the most common genes are *bla*_TEM_ and *bla*_CTX-M._

## Conclusion

The prevalence of UTI among refugees in Nakivale settlement is high with the commonest bacterial causes being *Staphylococcus aureus* and *Escherichia coli*. These organisms exhibit a very high rate of multidrug resistance to common drugs used to treat UTI with a high prevalence of ESBL-producing Enterobacteriaceae is high and the common ESBL genes are *bla*_TEM_ and *bla*_CTX-M._

## Data Availability

All relevant data are within the manuscript and its Supporting Information files.

NA

## Authors’ contributions

AHH conceptualized the study and participated in data collection. KK contributed to the design and conduct of the molecular assays JM carried out the culture and sensitivity, KT guided the contributed to study design and analysis, BM contributed to data analysis, JB over saw the overall running of the study from start to finish and wrote the final manuscript.

All authors read and approved the final version of the manuscript.

